# Evaluation and demonstration of a new tuberculosis diagnostic tool for Indonesia: Study protocol of the EVIDENT cluster randomised controlled trial

**DOI:** 10.64898/2026.08.12.26360245

**Authors:** Nur Afifah, Raspati C. Koesoemadinata, Edwin Ardiansyah, Kurnia Wahyudi, Bony W. Lestari, Reinout van Crevel, Stephen Graham, Susan M. McAllister, Katrina Sharples, Philip C. Hill, Bachti Alisjahbana

## Abstract

**Introduction:** Most people with tuberculosis (TB) reside in countries with limited resources for prompt TB diagnosis, resulting in diagnosis and treatment delays and ongoing community transmission. Current TB diagnostics rely on sputum, while some people with presumptive TB cannot produce adequate sample. A new generation near-point-of-care (NPOC) tests using sputum or tongue swabs may provide more accessible TB diagnosis.

**Methods:** In this pragmatic cluster randomised controlled trial (cRCT) in Indonesia, a multi-component public health intervention will include: (a) introduction of NPOC MiniDock MTB test (Guangzhou Pluslife Biotech, China) on sputum, tongue swab, or both specimens, (b) optimisation of clinical algorithms incorporating the new test, (c) a promotional package to encourage patient attendance and test utilisation, and (d) TB household contact investigation, including the new test, by community health volunteers. We will randomly stratify 40 community health centre (CHC) areas into intervention and control arms (1:1) in Bandung District. The intervention will be administered in healthcare facilities (HCFs) and in the community of the intervention areas. The control areas will continue standard of care with no intervention, except TB notification refresher training, which will be delivered in both areas before intervention roll-out. The primary outcome is the number of TB cases diagnosed and notified by HCFs per population attending them. It will be measured by abstracting data on TB case notification and the number of HCF attendees over 12 months following completion of the intervention roll-out, compared to 12 months preceding any trial activities.

**Discussion:** This trial will evaluate the effect of an intervention package incorporating the first-in-class NPOC on TB diagnosis and notification. The trial results will inform policy to improve TB diagnostic efforts in Indonesia and other high burden TB countries.

**Trial Registration:** ClinicalTrials.Gov, NCT07293455.

## Introduction

### Background and rationale

Tuberculosis (TB) remains a critical global health problem, with an estimated 10.7 million cases and nearly 1.23 million deaths annually.[1] Importantly, the majority of people with TB reside in low- and middle-income countries (LMIC), where resources are limited.[1] Moreover, a substantial proportion of individuals with TB in LMICs seek care in private health clinics or other peripheral clinics without adequate laboratory services to diagnose TB, resulting in diagnosis and treatment delays.[2] These delays not only affect an individual’s TB prognosis but also drive *M. tuberculosis* transmission in the community.

Early diagnosis and high treatment coverage are critical for effective TB control. However, even with the roll-out of WHO-approved rapid molecular diagnostics for TB, access to testing remains largely concentrated at centralised, government-owned healthcare facilities (HCFs) equipped with medium to high-capacity laboratories. Satellite or smaller HCFs with inadequate laboratory capacity commonly rely on less sensitive diagnostic tests like sputum microscopy for acid-fast bacilli (AFB) or referral of presumptive TB patients to other facilities.[2,3]

Furthermore, most diagnostics for microbiological confirmation are dependent on the quality of the sample provided for laboratory evaluation. Approximately a fourth of presumptive TB patients cannot produce an adequate sputum sample.[4] Therefore, there is an urgent need to develop and implement new rapid diagnostic tests on alternative samples that are scalable and easily adaptable for use in resource-limited settings.[5] Using new-generation diagnostic testing methods on tongue swabs provides such an opportunity.

In Indonesia, which holds approximately 10% of the global TB burden, the national health system is complex.[1] TB care and management are primarily delivered in community health centres (CHC) and hospitals. However, it is also common for individuals to attend pharmacies and smaller private clinics, which lack diagnostic services,[2] while Gene Xpert testing is only available in some CHCs and often facing operational challenges[3]. This paper describes a protocol of the EVIDENT cluster randomised controlled trial (cRCT) to evaluate a diagnostic intervention package based around a new near point-of-care (NPOC) molecular diagnostic test, MiniDock MTB test (Guangzhou Pluslife Biotech, China) on sputum or tongue swabs, including optimisation of clinical algorithms around the new test, promotional packages to encourage attendance for testing, and TB contact investigation. This trial will provide evidence to inform national TB diagnosis guidelines and optimal pathways to incorporate the new diagnostic test.

### Objectives

The primary objective of the trial is to evaluate whether the diagnostic intervention package increases diagnosis and notifications of TB cases from HCF per population attending in comparison to control areas.

Secondary objectives for the trial are to evaluate whether:

i. the diagnostic intervention package will increase the number of TB cases notified in CHC areas, for people who live in the area, per CHC area population in comparison to control areas;
ii. the proportion of cases diagnosed with TB that are microbiologically confirmed will be higher in the HCF in the intervention areas than in the control areas;
iii. the time to TB diagnosis from the first visit to a formal HCF will be lower in the intervention than control areas;
iv. the number of visits to HCFs prior to TB diagnosis will be lower in the intervention than control areas;
v. the propensity to test for TB will be higher amongst HCFs in the intervention than in control areas;

and to calculate and compare:

vi) patient costs for TB diagnosis among patients diagnosed by the HCFs in intervention and control areas;
vii) TB diagnostic costs in the HCFs and community settings in intervention and control areas.

The primary and the first two secondary objectives will be measured in the main study (implementation of diagnostic intervention package) and are described in this manuscript. Other objectives will be measured in sub-studies alongside the trial which will be described separately.

## Materials and Methods

### Study Design

This is a cRCT of a multi-component public health intervention to increase the diagnosis and notification of TB cases in Bandung District (registered at ClinicalTrials.Gov; NCT07293455). Trial clusters are CHC areas, defined as geographical areas surrounding a CHC. The intervention will be administered in HCFs and in the community of the intervention CHC areas.

The control areas will continue the standard of care with no intervention, except a refresher training on reporting and notifying TB cases to the SITB, which will be delivered to all HCFs in both arms before the intervention package is rolled out. **Fig 1** summarises the timeline of study enrolment, intervention, and assessments in this trial. The development of this trial protocol adhered to the Standard Protocol Items: Recommendations for Interventional Trial (SPIRIT) 2025 guidelines (**S1 Checklist, S2 File, and S3 File**).[6]

**Fig 1.**
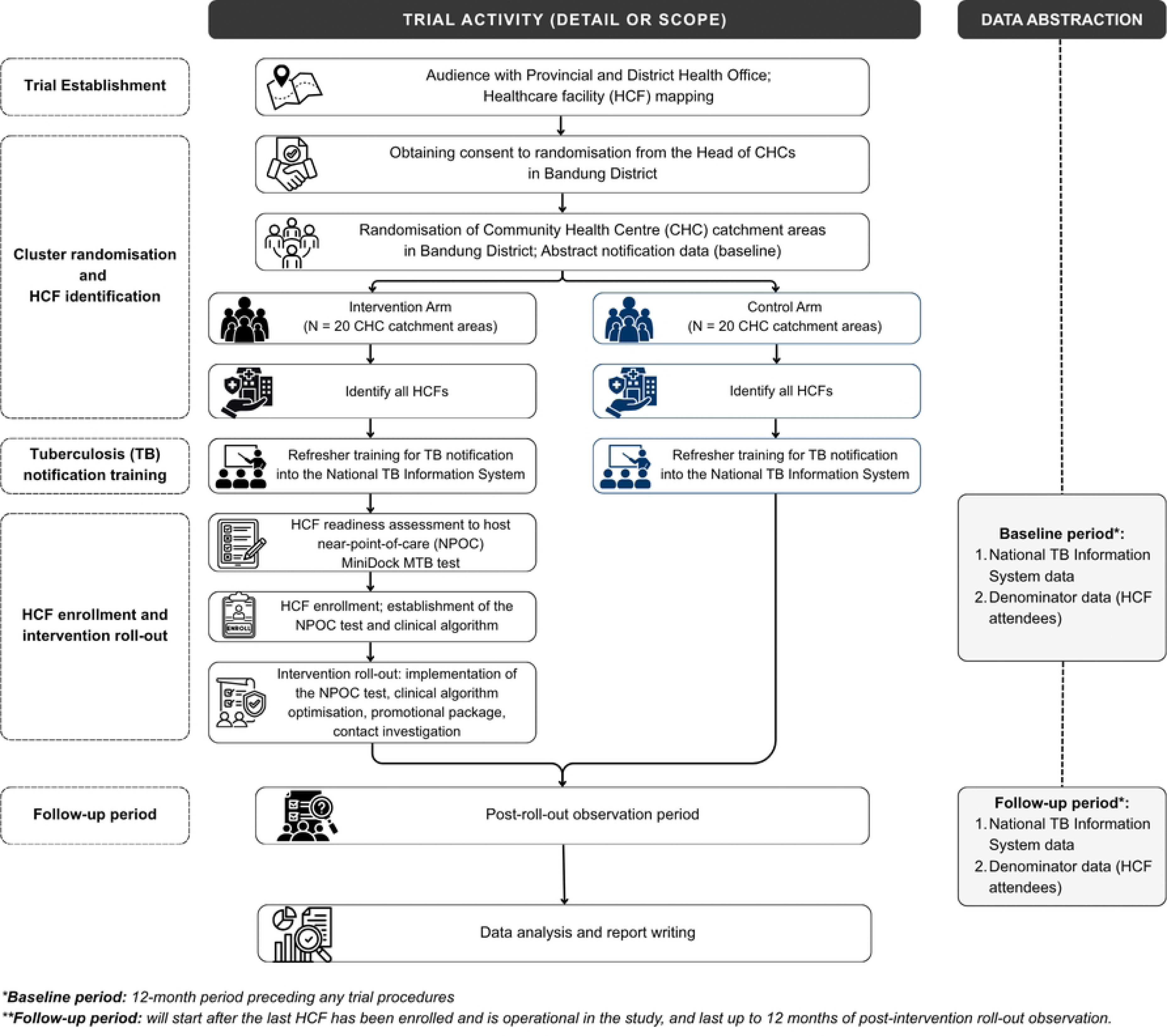
SPIRIT timeline for study enrolment, intervention, and assessments for the EVIDENT trial. CHC, community health centre; HCF, healthcare facility; NPOC, near point-of-care; TB, tuberculosis.

The trial schema will occur with the following components (**Fig 2**):

- Trial establishment: We will set up the site for the cRCT by having an audience with the Health Offices and conducting HCF mapping in all CHC catchment areas in Bandung District.
- Cluster randomisation and HCF identification: We will obtain group-level consent for randomisation from the heads of CHCs in Bandung District, perform cluster randomisation, and identify HCFs in the study area.
- TB notification training: All study HCF personnel (intervention and control) will receive refresher training on TB notification into the National TB Information System to maximise completeness of TB notification across all HCFs in both arms.
- HCF enrolment and intervention roll-out: A finalised intervention package will be implemented in the intervention areas.
- Follow-up period: After the last HCF has been enrolled and is operational in the intervention arm, the follow-up period will last up to 12 months of post-intervention roll-out.
- End of the study: The end of the period of analysis and write-up.

**Fig 2.**
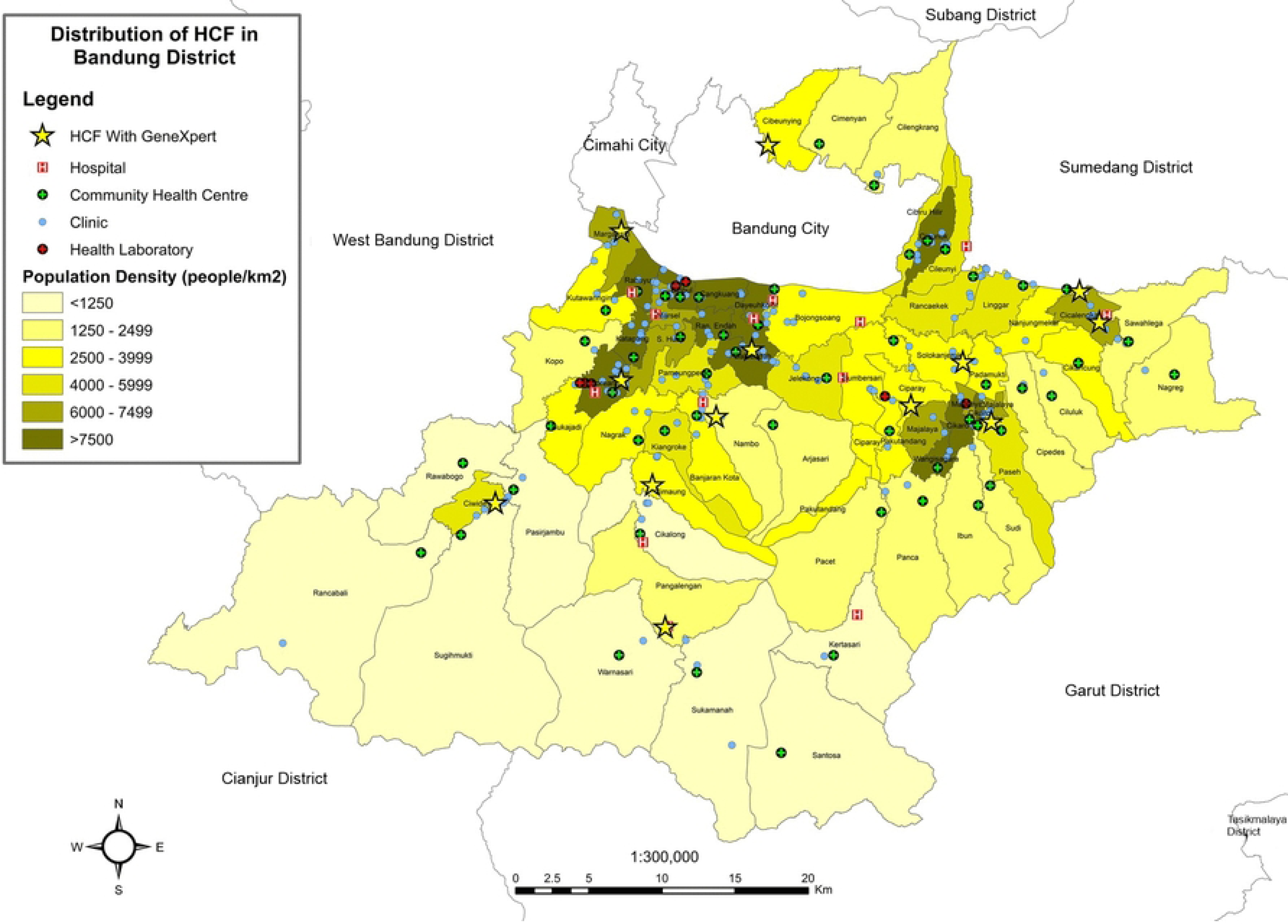
Schema of the EVIDENT trial. CHC, community health centre; HCF, healthcare facility; NPOC, near point-of-care; TB, tuberculosis.

### Setting

The study will be conducted in Bandung District, West Java Province, in Indonesia. Bandung District has an area of approximately 1,700 km^2^ with over 3.5 million inhabitants, giving a population density of approximately 2000 people/km^2^.[7] At the primary level, the District is served by 62 publicly-funded CHCs (each serving a defined ‘CHC catchment area’), and approximately 190 General Practitioner (GP) clinics, most of which are privately owned. At the secondary and tertiary referral levels, there are 15 private specialist clinics and 11 general hospitals. There are nine health laboratories in the District.[7]

Before developing the trial protocol, preliminary data collection was conducted to map the latest distribution of HCFs in the District (**Fig 3; S4 Table**) and to prepare for a subsequent simulation study. Our data indicated that the number of TB notifications and the proportion of TB notifications for patients living in each CHC area vary. In 2023, a total of 12,003 TB cases were notified to the National TB Information System (locally known as *Sistem Informasi Tuberkulosis* or *SITB*) from the District of which 3,661 were bacteriologically confirmed cases and 8,341 were clinically diagnosed.[8] Most private clinics do not have TB diagnostic facilities and therefore refer presumptive TB patients to another HCF. Moreover, only about a third (n=63) of the estimated 177 private clinics in the District have created accounts in the SITB for their clinics to notify TB cases.[9]

**Fig 3.**
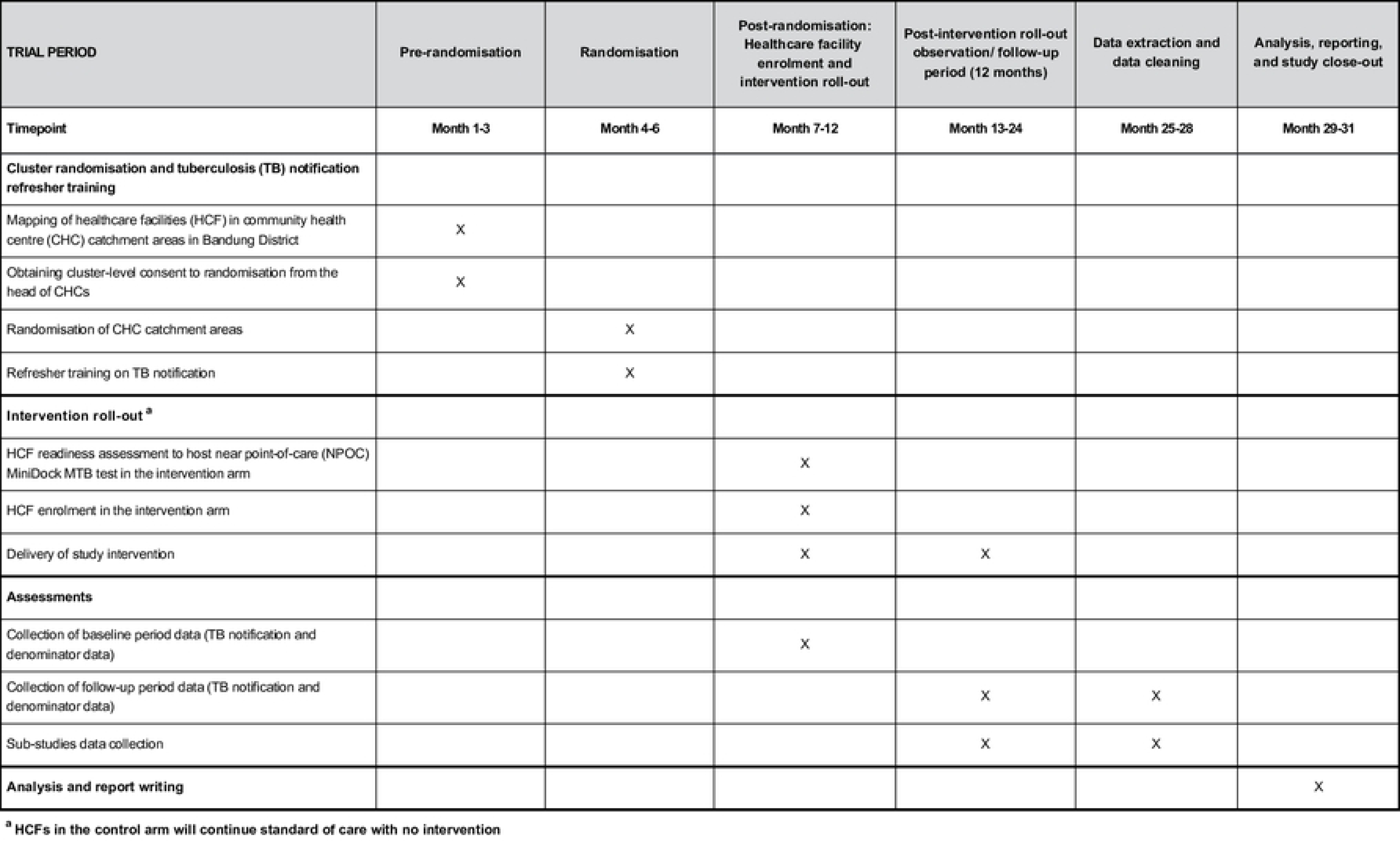
Distribution of healthcare facilities (HCF) in Bandung District.

### Randomisation and blinding

All CHC areas in Bandung District, except for one CHC with a large referral hospital and one other CHC due to the involvement in the ‘One Stop Service for TB’ program conducted by the Ministry of Health, are eligible for inclusion in the study (N=60). A random sample of 40 will be selected for the trial with stratification by whether or not Xpert testing is available in the CHC area and the background TB notification numbers. CHC areas with more than 400 TB notifications will be treated as a single stratum due to the small numbers (**Table 1**). All HCFs within the selected CHC areas are eligible for inclusion.

**Table 1.** Number of CHC catchment areas by availability of Xpert testing and number of TB notifications.

| Number of TB notifications (2023) | Xpert TB testing available in the CHC catchment area in Bandung District |  | Total |
| --- | --- | --- | --- |
|  | No | Yes |  |
| >400 | 1 | 4 | 5 |
| 84- 400 | 23 | 4 | 27 |
| <84 | 25 | 3 | 28 |
| Total | 49 | 12 | 60 |

CHC, community health centre; TB, tuberculosis.

The 40 selected CHC areas will be randomised 1:1 using the same strata as the selection of CHC areas. Both the selection and randomisation will be carried out independently by the trial overseas statistician (KS), who is unfamiliar with the CHC areas. Enumerators who will collect denominator data and person who will conduct data analysis will be blinded to randomisation allocation.

### Intervention

The intervention will have four components:

1. Introduction of a new-generation diagnostic test (MiniDock MTB test) using sputum, tongue swab, or both specimens, with a testing and clinical algorithm. If the patients can produce adequate-quality sputum, the NPOC test will be performed using a sputum swab. Otherwise, a tongue swab specimen will be collected on the same day, and the NPOC test will be performed using both specimens. For those who cannot or decline to produce sputum, tongue swab testing will be done. Staff in intervention HCFs will receive training on the new tests from sample preparation through to data recording and notification.
2. Optimisation of clinical algorithms with the incorporation of the new test will be co-designed with representative clinical staff from facilities. The new algorithms will take into account: risk factors for multidrug-resistant TB, the availability of Xpert Ultra testing and chest X-rays at each facility, and how these can be optimally used in conjunction with the new test. Clinical decisions regarding the establishment of TB diagnosis and the initiation of TB treatment will be provided at each HCF, in accordance with National TB Guidelines.
3. Promotional package to encourage patient attendance and utilisation of NPOC tests at HCFs. A large number of TB patients visit a pharmacy when they first experience symptoms.[2] To increase attendance of presumptive TB patients at HCFs, pharmacy staff and healthcare workers at HCF not hosting the test in the intervention areas will be trained to undertake a simple symptom screen and refer to the nearest HCF for TB testing as appropriate. They will also receive educational material on the signs and symptoms of TB and TB diagnosis, including information about the new tests.
4. TB household contact investigation by community health volunteers (cadres) who live in the area and provide routine support for TB patients. Cadres will be trained to perform symptom screening of household contacts, collect specimens, and transport them. Specimen collection will be performed following the above-mentioned specimen triage process. Specimen collection will either occur at the HCF connected to the index case or on-site at the household. Further management of household contacts will be determined by the clinicians, according to National TB Guidelines.

Leaders from all eligible HCFs in the intervention areas will be invited to a seminar and/or approached at their workplaces to receive information about the study, the new-generation TB test, and the incorporation of the test into routine practice using adjusted clinical algorithms. Each HCF will be visited and assessed using a standardised HCF assessment tool to ensure suitability for hosting the new TB diagnostic test. Written informed consent will be obtained from each HCF. For each component of the intervention, monitoring of delivery and uptake will be assessed according to performance indicators on a monthly basis.

### Outcome measures

The primary outcome measure, at the HCF level, is the number of TB cases diagnosed and notified by each HCF per population attending them. Secondary outcome measures in the main study include: the number of TB cases diagnosed and notified from individual CHC areas, who live in the area, per CHC area population; and the number of diagnosed and notified TB cases that are microbiologically confirmed over the number of TB cases diagnosed in each HCF.

Primary and secondary outcomes in the main study will be assessed over a 12-month period following completion of the intervention roll-out, which will occur over a maximum time of 6 months. Baseline data for the 12-month period preceding any trial procedures will also be collected.

### Data collection and management

The number of TB cases notified in all study CHC areas will be assessed by abstracting routinely recorded notifications for the baseline and follow-up periods (**Table 2**). Data abstracted will include notifying HCF and CHC, date of notification, patient’s demographic information (national identification number and name to check for duplicates, age, birth date, sex, residential address, and occupation), previous TB history, results of investigations and clinical assessments (paediatric TB scoring; chest X-ray; sputum microscopy, Xpert and/or culture; and other diagnostic test results), the basis of TB diagnosis (clinical diagnosis or bacteriological confirmation), location of TB disease (pulmonary or extrapulmonary), comorbidity status, information on contact investigation, TB treatment start date, and TB treatment outcomes. The CHC of residence for each notified patient will be matched to their residential address.

**Table 2.** Data sources for TB notifications and denominator data.

| No. | Data | Source |
| --- | --- | --- |
| 1 | TB cases notified | National TB Information System |
| 2 | TB cases diagnosed but not notified | HCF data |
| 3 | People attending HCFs | HCF visit, District Health Office<br>(Department of Healthcare) |
| 4 | CHC area population | District Health Office, National Statistics<br>Office of Indonesia |

CHC, community health centre; HCF, healthcare facility; TB, tuberculosis.

Denominator data will include population data obtained through Bandung District Health Office and the National Statistics Office of Indonesia, and the number of people attending HCFs obtained by visiting all HCFs in the intervention and control arms. This will be supplemented by data from the Department of Healthcare of the District Health Office (**Table 2**).

The first data extraction will be carried out three months after the start of intervention roll-out to allow time for data maturity and cleaning by the National TB Information System and District Health Office. An interim analysis will be carried out following this extraction to test the data extraction processes and data quality, and to set up the final analyses. Extraction of TB notification data will be conducted every three months after the first data abstraction, through to the end of the follow-up period. All data extractions will be carried out in collaboration with the District Health Office, Provincial Health Office, and the National Statistics Office of Indonesia. Trained enumerators, blinded to the treatment allocation, will abstract data on the number of HCF attendees. We will coordinate with the District Health Office for data collection.

Visits will be made to all HCFs in both intervention and control arms at the beginning of the post-roll-out observation period and after six months to monitor completeness of notifications and adherence to the diagnostic algorithms according to pre-determined performance indicators and a standard form to record results. Every effort will be made to keep all HCFs in the trial to minimise bias, however, any HCF that is unable to meet the test hosting requirements consistently will be withdrawn by the trial PI, and any HCF that chooses to withdraw will be free to do so. Notifications from these facilities will continue to be included in the study data abstraction of routinely collected data from the SITB.

Retention of all HCFs in the study is important for collection of denominator data on numbers of HCF attendees. HCF who cannot host the intervention, do not wish to take part in the intervention or wish to withdraw from the intervention will be encouraged to remain in the study for the purposes of collecting denominator data.

### Statistical analysis

Analysis for the primary objective (HCF notifications) will estimate the population average treatment effect as a rate ratio adjusted for baseline notification rates and stratification factors using a constrained baseline log-linear longitudinal data model with Poisson errors with a generalised estimating equation to provide robust standard errors accounting for both overdispersion and clustering by CHC areas.[10] The estimated absolute difference in notification rates per population visiting HCFs between intervention and control areas will be provided with a 95% bootstrap confidence interval. Sensitivity analyses to explore the effect of any under-notification on the comparison of intervention and control arms. Analysis for secondary objective 1 will follow the same approach as for the primary objective with CHC areas as the unit of analysis. For secondary objective 2, the analysis will estimate an odds ratio comparing the odds of microbiological confirmation in notified TB cases in intervention and control arms using a generalised estimating equation (GEE) framework with a logistic link and binomial errors, clustering by CHC area and robust standard errors. The model will include terms for the treatment arm and the stratification. A further analysis will compare HCFs with and without the new test in the intervention group to the control HCFs.

### Sample size justification

We carried out a simulation study based on the 2023 TB notifications and estimated HCF attendees. Briefly, we used the 2023 number of TB notifications at each cluster, divided into subgroups to represent numbers of HCF notifications, to generate TB notifications at baseline and follow-up using a negative binomial model with a dispersion parameter of 1.2. The number of replications in the simulation was 3,000. We explored various treatment effects (rate ratios) of 1.2, 1.3 and 1.4, and varying duration of follow-up (9 months and 12 months). A true rate ratio of 1.3 was considered the minimum improvement in notifications rates that would make the intervention programme worthwhile. With 20 clusters per arm and 12 months follow-up if the true rate ratio was 1.3 for the constrained longitudinal data model, the power was estimated to be 90%, with significance set at the 5% level (two-sided) (**S5 Table**).

### Interim Analyses

This trial will involve two interim analyses. The first interim analysis will be conducted after baseline data collection to evaluate the integrity of outcome data and trial conduct, without any formal statistical analyses. Corrective actions will be taken before the follow-up period as part of quality control measures, enabling early issue detection without impacting the final outcome analysis.

The second interim analysis will be conducted six months after the start of the follow-up period. It will will focus on monitoring early trends of intervention implementation and assessing patient pathways. We will use the findings for internal quality purposes and will not involve formal hypothesis testing or early stopping decisions.

### Patient and public involvement

Engagement with key stakeholders at the local study sites, such as the Provincial Health Office, District Health Office, and CHCs, will be carried out at the beginning and throughout the project to align with the National TB Programme and global agencies.

### Safety considerations

Staff and cadres involved in specimen collection, transport, and testing may be at risk of exposure to *M. tuberculosis*. This risk will be minimised through training and adherence to study standard operating procedures (SOPs) for specimen collection, handling, testing, transport, and waste disposal. Healthcare workers and cadres performing diagnostic procedure will use appropriate personal protective equipment (PPE), including N95 masks, gloves, and gowns.

### Ethics and informed consent

The study has received initial ethical approval from the Research Ethics Committee, Universitas Padjadjaran Bandung (Number 467/UN6.KEP/EC/2025) on May 21, 2025. A protocol amendment was reviewed and approved by the same committee (Number 7/UN6.KEP/EC/2026) on February 5, 2026, which included modifications to the information sheet and consent forms for the NPOC test to support the pragmatic aspects of this trial.

All eligible HCFs in the intervention areas will be approached for their consent to participate in the intervention. Each participant will be asked to provide written informed consent, and their associated data will be identified using a unique study identification number. Study data systems will be password protected, and all study-related documentation will be securely stored and treated confidentially. At the conclusion of the study, de-identified databases will be archived in accordance with ethical, institutional, and sponsor requirements. De-identified data may be used for future research only after appropriate permissions, ethical approval, and data-use agreements have been obtained.

Study oversight will be provided by a Trial Steering Committee. Day-to-day operations will be managed by a Trial Management Group, and a data monitoring will be conducted by a Data Monitoring Committee; both will report to the Trial Steering Committee.

### Trial Status

As of this submission, the trial is actively progressing in accordance with Protocol version 2.0, dated November 18, 2025. TB notification refresher training was conducted on HCFs in the intervention and control arms in August 2025. HCF enrolment and intervention roll-out were conducted between September 2025 and February 2026. The delivery of the diagnostic intervention package is currently ongoing and expected to conclude by March 2027. Formal statistical analysis for the trial objectives will begin after the end of the follow-up period.

## Discussion

In February 2026, the World Health Organisation (WHO) recommended the use of the NPOC molecular test for TB diagnosis.[11] MiniDock MTB test is the first-in-class NPOC test which can examine swabs of sputum or tongue.[11,12] Previously, several cRCTs were conducted to evaluate the use of NPOC Molbio Truenat MTB-Plus and Xpert MTB/RIF in the primary care level; however, they relied on sputum specimens.[13–15] To our knowledge, the EVIDENT trial is the first cRCT to evaluate the effect of a multi-component public health intervention package using the NPOC MiniDock MTB test on TB diagnosis and notification at HCF and community settings. This trial answers a call to action to the national government, donors and global health actors, and civil society to ambitious NPOC test roll-out into the health system.[16]

The NPOC test implementation will be integrated within a multi-component intervention package, while in parallel, the control areas will continue standard of care. Therefore, the primary analysis will aim to evaluate the effectiveness of the package rather than the independent effect of its components. To supplement information, we will conduct several sub-studies to evaluate the patient pathway, the propensity to test, and the cost of the individual intervention at the HCF and community levels.

The intervention package will be delivered to the health system in a district setting in Indonesia, engaging both public and private HCFs. We will involve various types of formal HCFs, except those whose healthcare services provided by a single physician (‘solo practices’) which are mostly not equipped with in-house laboratories. Furthermore, we will strengthen the external linkage by encouraging referral of patients with TB symptoms from private clinics that are not eligible to host the NPOC test and also pharmacies near the HCFs that are eligible to host the NPOC test.

At the time of intervention roll-out, the NPOC MiniDock MTB test is a new diagnostic tool that has not been used in the Indonesian health system. Thus, we expect the degree of contamination between study arms in relation to test availability at facilities to be minimal. However, we have noted that individual treatment-seeking behaviour among presumptive TB patients is often not restricted to the area where people live.[17] Hence contamination in relation patients attending HCFs in a different trial arm is likely to be substantial, which resulted in the decision for the primary endpoint to be focused on a change in TB notifications from the HCFs themselves in the intervention areas, rather than utilising cluster area populations for denominators.

Given the pragmatic nature of the trial, blinding of treatment allocation for study investigators and research assistants is impractical. However, the enumerators will be blinded to minimise potential bias in the collection of outcome data, as will those conducting the analysis. Allocation concealment for the Provincial Health Office, District Health Office, and CHCs is not possible, as they are in regular communication regarding the TB program. TB notifications from all HCFs in both study areas will be obtained from the District Health Office. Although we will perform refresher trainings on TB notification for these HCFs before rolling out the intervention package, underreporting is likely, particularly among private HCFs.[18]

Before the intervention roll-out, we will conduct a standard assessment to determine HCF eligibility to host the NPOC MiniDock MTB test. Not all private clinics are expected to be eligible to host the test due to limitations in infrastructure and resources for in-house laboratory testing, which may mean that the true beneficial effect of the intervention is underestimated.[2] To maximise the uptake of the intervention, we will provide training and SOPs to healthcare providers in eligible HCFs and cadres within the intervention areas. The study training for HCFs and cadres will be conducted in collaboration with the Provincial and District Health Office, which may lead to greater engagement with the TB program among HCFs in the intervention areas.

In conclusion, this trial, along with the sub-studies, will provide valuable evidence on whether the multi-component public health intervention package is effective in increasing the number of TB diagnoses by measuring routine TB notifications. The study results will inform policy to improve TB diagnosis efforts in Indonesia and other similar settings.

## Authors’ contributions

**Conceptualization:** Nur Afifah, Raspati Cundarani Koesoemadinata, Reinout van Crevel, Stephen Graham, Katrina Sharples, Philip Campbell Hill, Bachti Alisjahbana

**Formal analysis:** Edwin Ardiansyah, Kurnia Wahyudi, Katrina Sharples

**Funding acquisition:** Raspati Cundarani Koesoemadinata, Philip Campbell Hill, Bachti Alisjahbana

**Investigation:** Nur Afifah, Raspati Cundarani Koesoemadinata

**Methodology:** Edwin Ardiansyah, Kurnia Wahyudi, Katrina Sharples, Philip Campbell Hill, Bachti Alisjahbana

**Project administration:** Nur Afifah, Raspati Cundarani Koesoemadinata, Bony Wiem Lestari, Susan Margaret McAllister, Bachti Alisjahbana

**Supervision:** Nur Afifah, Raspati Cundarani Koesoemadinata, Bony Wiem Lestari, Reinout van Crevel, Stephen Graham, Susan Margaret McAllister, Philip Campbell Hill, Bachti Alisjahbana

**Visualization:** Nur Afifah, Raspati Cundarani Koesoemadinata

**Writing – Original Draft Preparation:** Nur Afifah, Raspati Cundarani Koesoemadinata

**Writing – Review & Editing:** Nur Afifah, Raspati Cundarani Koesoemadinata, Edwin Ardiansyah, Kurnia Wahyudi, Bony Wiem Lestari, Reinout van Crevel, Stephen Graham, Susan Margaret McAllister, Katrina Sharples, Philip Campbell Hill, Bachti Alisjahbana

## Data Availability

No datasets were generated or analysed during the current study. All relevant data from this study will be made available upon study completion.

## Acknowledgements

We would like to thank the Bandung District Health Office and the Head of HCFs for their support in carrying out this trial. We would also like to thank Lubabul Aniq and Balqist Kharisma Nayu for assisting with the development of the figures (map of HCF distribution in Bandung District and trial schema figure).

## Supporting information

**S1 Checklist. The SPIRIT 2025 cheklist. S2 File. Approved trial protocol (English)**

**S3 File. Approved trial protocol (Original)**

**S4 Table. Preliminary data for each community health centre (CHC) catchment area.**

**S5 Table. Power for the analysis with 9 and 12-months duration of follow-up, 40, 50 and 60 CHC clusters (total), rate ratios of 1.2, 1.3 and 1.4, and a two-sided Type I error rate of 5%.**

## References

1. World Health Organization. Global tuberculosis report 2025. Geneva: WHO; 2025.

2. Lestari BW, McAllister S, Hadisoemarto PF, Afifah N, Jani ID, Murray M, et al. Patient pathways and delays to diagnosis and treatment of tuberculosis in an urban setting in Indonesia. The Lancet Regional Health - Western Pacific. 2020;5. doi:10.1016/J.LANWPC.2020.100059

3. Nijman G, Alifia A, Annisa SN, Shurianto L, de Boer IE, Puspitasari I, et al. Operational performance of GeneXpert for tuberculosis diagnosis in West Java province, Indonesia: a public health evaluation. BMC Health Services Research. 2026. doi:10.1186/s12913-026-14658-0

4. Sputum scarcity among adolescents and adults with presumptive tuberculosis: a systematic review and meta-analysis | medRxiv [Internet]. [cited 2026 Jun 24]. Available from: https://www.medrxiv.org/content/10.1101/2025.11.02.25339326v1.full

5. Pai M, Dewan PK, Swaminathan S. Transforming tuberculosis diagnosis. Nature Microbiology. 2023;8:756–9. doi:10.1038/s41564-023-01365-3

6. Chan AW, Boutron I, Hopewell S, Moher D, Schulz KF, Collins GS, et al. SPIRIT 2025 statement: Updated guideline for protocols of randomised trials. PLOS Medicine. 2025;22:e1004589. doi:10.1371/journal.pmed.1004589

7. Bandung District Health Office. Bandung District Health Profile 2022. Bandung; 2023.

8. Bandung District Health Office. Tuberculosis notification data [Unpublished raw data]. National Tuberculosis Information System. 2023.

9. The Ministry of Health of the Republic of Indonesia. Indonesian tuberculosis data portal [Internet]. Available from: https://data.sitb.id/fasyankes/v_fasyankes_list.php?goto=467

10. Hooper R, Forbes A, Hemming K, Takeda A, Beresford L. Analysis of cluster randomised trials with an assessment of outcome at baseline. BMJ. 2018;360:k1121. doi:10.1136/bmj.k1121

11. World Health Organization. Near point-of-care nucleic acid amplification tests (NPOC-NAATs) as a new diagnostic class for diagnosis of TB using sputum and tongue swabs [Internet]. Available from: https://www.who.int/teams/global-programme-on-tuberculosis-and-lung-health/diagnosis-treatment/npoc-tongue-swabs-and-sputum-pooling-for-tb/npoc-naats

12. Steadman A, Kumar KM, Asege L, Kato-Maeda M, Mukwatamundu J, Shah K, et al. Diagnostic accuracy of swab-based molecular tests for tuberculosis using near-point-of-care platforms: a multi-country evaluation. eBioMedicine. 2025;121. doi:10.1016/j.ebiom.2025.105991 PubMed PMID: 41175672.

13. Khosa C, Cossa M, Leukes V, Hella J, Sabi I, Watson M, et al. Implementing the Molbio Truenat platform and tuberculosis assays versus standard of care at primary care clinics for the detection and treatment of tuberculosis in Mozambique and Tanzania (TB-CAPT CORE): a cluster-randomised trial. The Lancet Primary Care. 2025;1. doi:10.1016/j.lanprc.2025.100028

14. Cattamanchi Adithya, Reza Tania F., Nalugwa Talemwa, Adams Katherine, Nantale Mariam, Oyuku Denis, et al. Multicomponent Strategy with Decentralized Molecular Testing for Tuberculosis. New England Journal of Medicine. 2021;385:2441–50. doi:10.1056/NEJMoa2105470

15. Lessells RJ, Cooke GS, McGrath N, Nicol MP, Newell ML, Godfrey-Faussett P. Impact of Point-of-Care Xpert MTB/RIF on Tuberculosis Treatment Initiation. A Cluster-randomized Trial. Am J Respir Crit Care Med. 2017;196:901–10. doi:10.1164/rccm.201702-0278OC PubMed PMID: 28727491; PubMed Central PMCID: PMC5649979.

16. Lynch S, Mupfumi L, Otieno C, Sharma A, Mataka A. The case for ambition: Why countries must move boldly on Near Point-of-Care TB Diagnostics. PLOS Global Public Health. 2026;6:e0006134. doi:10.1371/journal.pgph.0006134

17. Hadisoemarto PF. Tuberculosis case management and notification by private practitioners in Indonesia [University of Otago]. 2024. Available from: https://ourarchive.otago.ac.nz/esploro/outputs/doctoral/Tuberculosis-case-management-and-notification-by/9926619179701891#file-0

18. World Health Organization. The second national TB inventory study in Indonesia [Internet]. [cited 2026 Apr 7]. Available from: https://www.who.int/teams/global-programme-on-tuberculosis-and-lung-health/tb-reports/global-tuberculosis-report-2024/featured-topics/the-second-national-tb-inventory-study-in-indonesia

